# The impact of quality of primary care on secondary healthcare utilisation for patients with multiple long-term conditions

**DOI:** 10.64898/2026.08.13.26358683

**Authors:** Qian Gao, Benedict Hayhoe, Meryem Cicek, Geva Greenfield, Michaela Otis, Gesthimani Misirli, Ana Luisa Neves, Azeem Majeed, Paul Aylin, Alex Bottle

## Abstract

**Objectives:** To assess the concurrent and lagged associations between quality of primary care and planned and unplanned secondary care use for patients with multimorbidity, examining the modifying role of frailty.

**Design:** A retrospective cohort study

**Setting:** This population-level analysis included 468,172 patients with multimorbidity in England from the Discover research platform (April 2022-March 2024).

**Participants:** Patients with multimorbidity

**Main outcome measures:** We used principal component analysis to combine a set of quality indicators (QIs) and assessed the impacts of QIs on both planned and unplanned care.

**Results:** Generally, patients with higher QI attainment also had higher likelihood of planned (outpatient visits) and unplanned care (emergency admissions and ED visits) utilisation. There was a lower lagged odds of elective hospital admissions in the following 12 months among those with higher attainment of multimorbidity-specific QIs (OR=0.94, 95%CI 0.93-0.95). In the complex multimorbidity cohort (≥3 conditions), multimorbidity-specific QIs were longitudinally associated with lower odds of elective admissions (OR=0.94, 95%CI 0.92-0.95) and outpatient visits (OR=0.96, 95%CI 0.95-0.98), while generic QIs were related to lower odds of outpatient non-attendance (OR=0.95, 95%CI 0.91-0.99). In non-frail patients with multimorbidity, multimorbidity-specific QIs were longitudinally associated with reduced odds of outpatient visits (OR=0.98, 95%CI 0.97-0.99), elective admissions (OR=0.92, 95%CI 0.90-0.94) and prolonged elective hospital stay (IRR=0.94, 95%CI 0.89-0.99).

**Conclusions:** Attainment of generic and multimorbidity QIs was generally associated with slightly increased planned and unplanned care. However, patients for whom we identified higher attainment of multimorbidity-specific QIs had lower odds of elective admissions and outpatient visits, especially for those with complex multimorbidity. Our research suggests that the quality of primary care may influence patients’ use of secondary care, with the potential to improve care for people with multimorbidity and warrant further investigation into management strategies.

## Background

Individuals living with multiple long-term conditions, often referred to as multimorbidity, have complex healthcare needs and use health services more frequently than those with a single condition. This increased rate of service utilisation in relation to multimorbidity is consistent across different types and levels of healthcare services ^1^. A previous systematic review found greater utilisation of primary and secondary healthcare among patients with multimorbidity than those without ^1^. Patients with multimorbidity often have higher rates of hospital admissions, unplanned readmissions, and longer stays ^1–3^, especially those with additional mental comorbidity such as depression ^4^, three or more co-existing chronic conditions, and those who are frail ^5^ or socially disadvantaged ^6^. Socioeconomic deprivation intensifies these challenges, with disadvantaged individuals more likely to develop multimorbidity at a younger age and to experience worse health outcomes ^7^.

Patients with multimorbidity frequently have increased needs for secondary care, and this association is amplified by frailty, which can complicate disease management and elevate the risk of adverse clinical outcomes ^8^. Those with frailty experience a decline in functional capacity and are more likely to be admitted and readmitted to hospital after discharge, stay for longer, and have a higher risk of mortality ^5,8,9^. However, evidence of associations between quality of care in the community or primary care setting and secondary care outcomes for patients with multimorbidity is limited, as is the impact of frailty on such associations.

Primary care plays a fundamental role in managing multimorbidity, with primary care clinicians familiar with dealing with the complexity and interconnections of multimorbidity. Appropriate and timely primary care can vastly improve recovery from poor health outcomes and long-term survival ^10^. Previous evidence supports the key role of high-quality primary care in supporting early diagnosis, patient referral and coordination of multimorbidity care ^11^, which can effectively reduce demands for unplanned secondary care services (e.g. emergency hospital admissions and department visits) ^12,13^. In the UK, general practitioners (GPs) are tasked with monitoring patients on a range of nationally set indicators across chronic conditions and their clinically relevant markers in a scheme known as the Quality Outcomes Framework (QOF) ^14^. QOF incentivises GPs to monitor specific chronic conditions but does not consider multimorbidity as a specific category of quality improvement. This may contribute to a more fragmented and condition-specific approach, limiting the effectiveness of primary care in supporting patients with multimorbidity.

This study aimed to explore the associations between quality of primary care, as measured by a range of selected quality indicators (QIs), and subsequent levels of secondary healthcare use in the short term among patients with multiple long-term conditions and examine the modifying role of frailty in the associations.

## Methods

### Data sources

We used the Discover-NOW Hub’s Whole Systems Integrated Care (WSIC) database, a local integrated care database of pseudonymised patient data in north-west London that captures data for a range of health services, including primary and secondary care, for approximately 95% of the population in north-west London. We identified an adult cohort (aged 18 years and above) with multimorbidity, registered with general practices from 1 April 2022 to 30 March 2023. Multimorbidity was defined as co-existence of two or more long-term conditions ^15^. Working with public partners, researchers, clinicians, and people with lived experience of LTCs, and referencing a curated list of long-term conditions outlined in the 2021/22 UK Quality and Outcomes Framework (QOF) ^14^ we co-produced a list of conditions: asthma, anxiety, atrial fibrillation, cancer, coronary heart disease (CHD) or ischaemic heart disease, chronic kidney disease (CKD), chronic obstructive pulmonary disease (COPD), dementia, depression, diabetes, epilepsy, heart failure, hypertension, schizophrenia, bipolar affective disorder and other psychoses, obesity, osteoporosis, peripheral arterial disease (PAD), rheumatoid arthritis, stroke/transient ischaemic attack (TIA) (Gao et al. 2026, *in press*). In the final cohort, we included those who had a diagnosis of two or more of these long-term conditions during the study period, based on SNOMED CT codes. We linked the health records of general practices and secondary care in the corresponding and following year for the multimorbidity cohort to generate study datasets.

### Measures

#### Outcomes: healthcare utilisation

We examined both planned and unplanned healthcare use within the same financial year of QI attainment (April 2022-March 2023) and in the year following QI attainment (April 2023-March 2024). For planned healthcare, we assessed some outcomes as binary (whether the patients had any outpatient visits, non-attendance for scheduled outpatient appointments, and any elective hospital admissions), and others as counts (the number of outpatient appointments and duration of hospital stays measured in days). Similarly, unplanned healthcare use was considered as binary (emergency hospital admissions and ED visits) and counts (length of emergency hospital stays in days and total time spent in ED visits in days within the year). Length of all hospital stays and total ED visit counts were right-censored at 365 days for patients due to the one-year observation period. We recorded same-day hospital discharges as lasting a half-day for analytical purposes.

### Exposure

#### Quality indicators (QI)

Based on a review of NICE guidelines ^16^, a clinically curated set of quality indicators (QIs) was identified: clinical activity relating to blood pressure, serum cholesterol, blood sugar levels, smoking, body mass index (BMI), annual influenza vaccination, alcohol consumption, polypharmacy reviews, frailty and functional assessment, reviewing the risk of hospital admission, review of care plans, mental health screening [depression/anxiety], and reviewing the risk of falls. Each indicator was defined using the SNOMED CT clinical code lists and recorded as met (yes vs no) annually.

### Covariates

We included patients’ demographic characteristics (age, sex, ethnicity [White, Asian, Black/African-Caribbean, mixed/other ethnic minorities]) and health-related confounders, such as numbers of long-term conditions, existence of polypharmacy (yes/no), and recorded electronic frailty index [eFI] in the last 12 months, (incorporating 36 health conditions components and categorised as fit (eFI 0-0.12) and frail (mild to severe frailty eFI>0.12)) ^17^, and area-level index of multiple deprivation (IMD) rank deciles (with the lowest decile representing the most deprived neighbourhoods).

### Statistical Analysis

To combine the multiple QIs, we first calculated correlations between individual QIs and checked the risk of multiple counting. We then conducted the KMO and Bartlett’s test of sphericity to test the correlation matrix before conducting factor analyses. Finally, we conducted principal components analysis (PCA) of the quality indicators, with a cut-off point for the loading values set at 0.3 to decide which variable is represented by each principal component (PC). We used negative binomial regression models to assess the concurrent and lagged associations between QI and amount of planned and unplanned healthcare utilisation (i.e. total number of emergency department visits, length of emergency/elective hospital stays, number of outpatient and inpatient appointments, number of hospital admissions) within the same and the following year. We applied logistic regression models for binary outcomes (i.e. outpatient attendance / non-attendance, elective / emergency hospital admission, and ED visits). We applied standardised principal components in modelling and adjusted for patients’ demographics (age, gender, ethnicity and IMD) and health-related factors (number of comorbidities) in all models. We then examined the role of frailty in modifying QI attainment and healthcare utilisation. In sensitivity analyses, we further explored the concurrent and lagged associations in different multimorbidity cohorts (patients with two morbidities and those with four or more morbidities). To balance the baseline measure of frailty status and mitigate confounders, we applied Inverse Probability Treatment Weighting (IPTW) between healthcare users and non-users (for planned and unplanned care) by weighting participants in the analysis by the inverse probability of receiving their actual treatment. We used IPTW-weighted negative binomial regression and logistic regression models.

## Results

The cohort comprised 468,172 patients with multimorbidity (*Table 1*). The mean age of participants was 56.5 years, with nearly half aged over 60 years. 54.6% were female, 46.5% were of White ethnicity, and half were in the lowest five deciles of deprivation. Half of the sample were mild, moderately or severely frail, with hypertension and diabetes the commonest conditions. 67.9% had two morbidities and 32.1% had three or more. In the year following QI attainment (*Supplementary Table 1*), Hospital use was common, particularly outpatient visits. The levels of healthcare use among patients with multimorbidity were slightly lower within the same year than in the following year.

**Table 1.** Characteristics of study samples.

| Characteristics | n (%) |
| --- | --- |
| **Registered patients** | 1,944,423 |
| Multimorbidity patients | 468,172 |
| **Age (Mean, SD)** | 56.5 (17.6) |
| < 30 years | 39,626 (8.5%) |
| 30-59 years | 214,080 (45.7%) |
| ≥60 years | 214,476 (45.8%) |
| **Gender** |  |
| Female | 255,758 (54.6%) |
| Male | 212,395 (45.4%) |
| Missing | 19 (<0.01%) |
| **Ethnicity** |  |
| White | 217,658 (46.5%) |
| Asian | 135,799 (29.0%) |
| Black/African-Caribbean | 52,848 (11.3%) |
| Mixed/others | 59,982 (12.8%) |
| Missing (inc. Not known/stated) | 1,885 (0.4%) |
| **Index of multiple Deprivation (Deciles)** |  |
| 1 (most deprived) | 13,356 (2.8%) |
| 2 | 11,312 (2.4%) |
| 3 | 56,987 (12.2%) |
| 4 | 73,764 (15.8%) |
| 5 | 79,481 (17.0%) |
| 6 | 70,118 (15.0%) |
| 7 | 59,145 (12.6%) |
| 8 | 40,569 (8.7%) |
| 9 | 37,125 (7.9%) |
| 10 | 25,363 (5.4%) |
| Missing | 1,052 (0.2%) |
| **Multimorbidity (median, IQR)** | 2 (2-3) |
| **Number of comorbidities (range)** | (2-11) |
| 2 morbidities | 317,778 (67.9%) |
| 3 morbidities | 89,987 (19.2%) |
| 4 or more morbidities | 60,407 (12.9%) |
| **Hypertension** | 215,120 (46.0%) |
| **Diabetes** | 125,374 (26.8%) |
| **Polypharmacy in past 12 months** |  |
| Yes | 158,066 (33.7%) |
| No | 294,770 (63.0%) |
| missing | 15,336 (3.3%) |
| **Frailty** |  |
| Fit | 198,337 (42.3%) |
| Frailty | 254,499 (54.4%) |
| Missing | 15,336 (3.3%) |

### Quality of primary care by demographic and health characteristics

Using PCA approach, we identified two PCs: PC1 ‘Generic QIs’ (which contained more blood pressure, BMI, alcohol consumption, serum cholesterol, annual influenza vaccination, blood sugar levels measurement) and PC2 ‘Multimorbidity-specific QIs’ (which contained more multimorbidity relevant care processes like medication reviews, review of care plans, frailty and functional assessment, reviewing risk of falls) (*Supplementary Tables 2-3; Supplementary Figure 1*). There were higher mean scores of principal components (generic QIs) for older people, males and ethnic minorities, as well as those with a greater number of comorbidities and frailty (*Table 2*). The level of multimorbidity-specific QI was higher among younger and older groups, females, individuals of White ethnicity, and those living with a greater number of comorbidities and frailty.

**Table 2.** PCA component scores (means/SD) by demographic and health characteristics.

| Characteristics |  | PC1 generic QIs |  | PC2 multimorbidity-specific QIs |  |
| --- | --- | --- | --- | --- | --- |
| **Age** |  |  |  |  |  |
|  | <30 years | -0.89 (0.87) |  | 0.17 (0.58) |  |
|  | 30-60 years | -0.20 (1.00) |  | -0.12 (0.78) |  |
|  | ≥60 years | 0.37 (0.84) | P<0.001 | 0.09 (1.22) | P<0.001 |
| **Gender** |  |  |  |  |  |
|  | Female | -0.04 (0.99) |  | 0.03 (1.01) |  |
|  | Male | 0.04 (1.01) | P<0.001 | -0.04 (0.98) | P<0.001 |
| **Ethnicity** |  |  |  |  |  |
|  | White | -0.09 (1.03) |  | 0.06 (1.01) |  |
|  | Asian | 0.20 (0.90) |  | -0.09 (1.02) |  |
|  | Black/African-Caribbean | 0.04 (0.99) |  | -0.04 (0.98) |  |
|  | Mixed/others | -0.11 (1.04) | P<0.001 | 0.01 (0.93) | P<0.001 |
| **Number of comorbidities** |  |  |  |  |  |
|  | 2 morbidities | -0.22 (0.99) |  | -0.07 (0.86) |  |
|  | 3 morbidities | 0.31 (0.87) |  | 0.02 (1.11) |  |
|  | 4 or more morbidities | 0.69 (0.75) | P<0.001 | 0.35 (1.36) | P<0.001 |
| **Frailty** |  |  |  |  |  |
|  | Fit | -0.39 (0.98) |  | -0.11 (0.71) |  |
|  | Frailty | 0.38 (0.84) | P<0.001 | 0.08 (1.19) | P<0.001 |

#### Main models

Higher compliance with generic and multimorbidity-specific QIs was associated with prolonged hospital stays concurrently and longitudinally (*Table 3*). In general, higher attainment of generic and multimorbidity-specific QIs was concurrently and longitudinally associated with increased likelihood of outpatient visits and longer elective hospital stays, as well as more emergency admissions and ED visits (*Tables 4-5*). Conversely, patients who had higher attainment of multimorbidity-specific QIs were less likely to have elective admissions than those with lower attainment of QIs. The concurrent associations were consistent with the lagged associations, though the effect sizes of QIs were slightly smaller.

**Table 3.** The concurrent and lagged effects of quality of primary care on hospital stays among patients with multimorbidity.

| Outcomes | Same year |  | Following year |  |
| --- | --- | --- | --- | --- |
|  | Length of hospital stay in single admissions (IRR, 95%CI) | Total length of hospital stay (IRR, 95%CI) | Length of hospital stay in single admissions (IRR, 95%CI) | Total length of hospital stay (IRR, 95%CI) |
| **All patients** |  |  |  |  |
| Generic QI | 1.34 (1.32-1.36) | 1.38 (1.35-1.40) | 1.22 (1.20-1.23) | 1.24 (1.22-1.26) |
| Multimorbidity-specific QI | 1.17 (1.15-1.18) | 1.19 (1.17-1.21) | 1.12 (1.11-1.14) | 1.15 (1.13-1.16) |
| **Frailty** |  |  |  |  |
| Generic QI | 1.12 (1.10-1.15) | 1.13 (1.11-1.16) | 1.05 (1.04-1.07) | 1.06 (1.04-1.08) |
| Multimorbidity-specific QI | 1.17 (1.15-1.18) | 1.20 (1.19-1.22) | 1.12 (1.11-1.13) | 1.15 (1.13-1.16) |
| **Non-frailty** |  |  |  |  |
| Generic QI | 1.38 (1.33-1.43) | 1.37 (1.32-1.43) | 1.18 (1.14-1.21) | 1.18 (1.14-1.22) |
| Multimorbidity-specific QI | 1.22 (1.17-1.28) | 1.20 (1.14-1.26) | 1.11 (1.06-1.16) | 1.13 (1.08-1.18) |

Note. Reference category lower QI attainment. OR: Odds ratios. IRR: incidence rate ratio.

**Table 4.** The concurrent effects of quality of primary care on healthcare utilisation among patients with multimorbidity.

| Outcomes | Planned |  |  |  |  | Unplanned |  |  |  |
| --- | --- | --- | --- | --- | --- | --- | --- | --- | --- |
|  | Outpatient visits<br>(OR,95%CI) | Outpatient non-attendance<br>(OR,95%CI) | Number of outpatient appointments<br>(IRR,95%CI) | Elective admissions<br>(OR,95%CI) | Length of elective hospital stay<br>(IRR,95%CI) | Emergency admissions<br>(OR,95%CI) | Length of emergency hospital stay<br>(IRR,95%CI) | ED visits<br>(OR,95%CI) | Total ED attendances<br>(IRR,95%CI) |
| **All patients** |  |  |  |  |  |  |  |  |  |
| Generic QI | 1.51 (1.50-1.52) | 1.06 (1.05-1.08) | 1.38 (1.37-1.39) | 1.27 (1.26-1.29) | 1.30 (1.28-1.33) | 1.33 (1.32-1.35) | 1.42 (1.40-1.45) | 1.31 (1.30-1.32) | 1.32 (1.31-1.33) |
| Multimorbidity QI | 1.06 (1.05-1.07) | 1.12 (1.10-1.13) | 1.06 (1.06-1.07) | 0.95 (0.94-0.95) | 1.06 (1.04-1.08) | 1.09 (1.09-1.10) | 1.23 (1.21-1.25) | 1.15 (1.14-1.16) | 1.17 (1.16-1.18) |
| **Frailty** |  |  |  |  |  |  |  |  |  |
| Generic QI | 1.37 (1.35-1.38) | 0.95 (0.93-0.97) | 1.19 (1.18-1.20) | 1.17 (1.16-1.19) | 1.14 (1.11-1.18) | 1.20 (1.19-1.22) | 1.17 (1.14-1.20) | 1.19 (1.18-1.21) | 1.18 (1.17-1.19) |
| Multimorbidity QI | 1.04 (1.03-1.04) | 1.13 (1.11-1.14) | 1.05 (1.05-1.06) | 0.95 (0.94-0.96) | 1.07 (1.05-1.09) | 1.10 (1.09-1.11) | 1.22 (1.20-1.24) | 1.15 (1.15-1.16) | 1.17 (1.16-1.18) |
| **Non-frailty** |  |  |  |  |  |  |  |  |  |
| Generic QI | 1.36 (1.35-1.38) | 1.07 (1.04-1.09) | 1.37 (1.35-1.39) | 1.17 (1.14-1.19) | 1.22 (1.16-1.28) | 1.26 (1.24-1.29) | 1.46 (1.39-1.52) | 1.25 (1.23-1.27) | 1.27 (1.25-1.29) |
| Multimorbidity QI | 1.02 (1.00-1.03) | 1.09 (1.05-1.12) | 1.16 (1.14-1.18) | 0.94 (0.92-0.97) | 1.08 (1.01-1.15) | 1.00 (0.98-1.02) | 1.31 (1.24-1.39) | 1.04 (1.02-1.06) | 1.10 (1.09-1.12) |

*Note.* Reference category lower QI attainment. OR: Odds ratios. IRR: incidence rate ratio.

**Table 5.** The lagged effects of quality of primary care on following 12-month healthcare utilisation among patients with multimorbidity.

| Outcomes | Planned |  |  |  |  | Unplanned |  |  |  |
| --- | --- | --- | --- | --- | --- | --- | --- | --- | --- |
|  | Outpatient visits<br>(OR, 95%CI) | Outpatient non-attendance<br>(OR, 95%CI) | Number of outpatient appointments<br>(IRR, 95%CI) | Elective admissions<br>(OR, 95%CI) | Length of elective hospital stay (IRR, 95%CI) | Emergency admissions<br>(OR, 95%CI) | Length of emergency hospital stay<br>(IRR, 95%CI) | ED visits<br>(OR,95%CI) | Total ED attendances<br>(IRR, 95%CI) |
| **All patients** |  |  |  |  |  |  |  |  |  |
| Generic QI | 1.24 (1.24-1.25) | 1.07 (1.06-1.08) | 1.18 (1.17-1.18) | 1.15 (1.14-1.16) | 1.14 (1.13-1.16) | 1.14 (1.14-1.15) | 1.15 (1.14-1.16) | 1.12 (1.11-1.12) | 1.12 (1.11-1.12) |
| Multimorbidity QI | 1.02 (1.01-1.02) | 1.08 (1.07-1.09) | 1.03 (1.02-1.03) | 0.94 (0.93-0.95) | 0.99 (0.97-1.00) | 1.07 (1.07-1.08) | 1.14 (1.12-1.15) | 1.09 (1.09-1.10) | 1.11 (1.10-1.11) |
| **Frailty** |  |  |  |  |  |  |  |  |  |
| Generic QI | 1.33 (1.32-1.35) | 0.98 (0.96-1.00) | 1.17 (1.16-1.18) | 1.18 (1.16-1.19) | 1.14 (1.10-1.17) | 1.14 (1.12-1.15) | 1.08 (1.06-1.11) | 1.09 (1.08-1.10) | 1.10 (1.09-1.11) |
| Multimorbidity QI | 1.00 (0.99-1.00) | 1.12 (1.11-1.13) | 1.03 (1.02-1.03) | 0.94 (0.93-0.94) | 1.00 (0.98-1.01) | 1.09 (1.08-1.10) | 1.16 (1.14-1.17) | 1.11 (1.10-1.12) | 1.12 (1.12-1.13) |
| **Non-frailty** |  |  |  |  |  |  |  |  |  |
| Generic QI | 1.33 (1.32-1.35) | 1.13 (1.11-1.16) | 1.31 (1.30-1.33) | 1.17 (1.15-1.20) | 1.13 (1.08-1.18) | 1.18 (1.16-1.21) | 1.22 (1.18-1.27) | 1.15 (1.14-1.16) | 1.16 (1.15-1.18) |
| Multimorbidity QI | 0.98 (0.97-0.99) | 1.04 (1.01-1.07) | 1.11 (1.09-1.12) | 0.92 (0.90-0.94) | 0.94 (0.89-0.99) | 0.99 (0.97-1.02) | 1.15 (1.09-1.21) | 1.02 (1.00-1.03) | 1.08 (1.07-1.10) |

There was some evidence that the association between attainment of QIs and planned healthcare use differed according to frailty. Longitudinally, high attainment of multimorbidity-specific QIs had slightly decreased odds of outpatient visits (OR=0.98, 95%CI 0.97-0.99), with a reduction in IRR for elective hospital stays (IRR=0.94, 95%CI 0.89-0.99) for non-frailty groups, but with no statistical differences for patients with frailty. Patients with higher attainment of multimorbidity-specific QIs had lower odds of elective hospital admissions (OR=0.94, 95%CI 0.93-0.95), with the associations persisting across frailty status (frail OR=0.94, 95%CI 0.93-0.94 and non-frail groups OR=0.92, 95%CI 0.90-0.94). We applied IPTW to better balance covariates across documented EFI measures (*Supplementary Table 4*), with IPTW-weighted and covariate-adjusted effect sizes consistent with the main models on all outcomes.

### Sensitivity analysis

The effects of QI attainment on secondary care utilisation were assessed across basic (two morbidities) and complex multimorbidity (four or more morbidities) cohorts in the sensitivity analysis. In both multimorbidity cohorts, higher attainment of generic and multimorbidity-specific QIs was associated with a higher number of outpatient appointments, elective and emergency hospital admissions, and ED visits after adjustment for sociodemographic characteristics (*Supplementary Tables 5-6*). Findings were generally consistent across multimorbidity cohorts, except for outpatient visits and non-attendance. In the basic multimorbidity cohort, high attainment of multimorbidity QIs was associated with slightly lower odds of elective admissions but with higher likelihood of other planned and unplanned care use. In the complex multimorbidity cohort, high attainment of multimorbidity QIs was associated with slightly lower odds of outpatient visits and elective admissions. Patients with high attainment of generic QI had lower odds of outpatient non-attendance. Compared with those with two or more comorbidities, effect sizes of QIs in planned and unplanned healthcare were slightly lower for complex multimorbidity.

## Discussion

### Summary of findings

This study provides evidence of associations between quality of primary care and a range of planned and unplanned healthcare utilisation in patients with multimorbidity. Higher attainment of both generic and multimorbidity-specific QIs was generally associated with higher odds of planned (outpatient visits) and unplanned care (emergency admissions and ED visits). In contrast, patients with higher attainment of multimorbidity-specific QIs had a reduced likelihood of elective hospital admissions. All effect sizes were modest. There was some evidence of a modifying role of frailty and complexity of multimorbidity in the associations between attainment of QIs and planned healthcare use. Although higher attainment of generic and multimorbidity-specific QIs was generally associated with increased outpatient utilisation, this pattern differed in those with greater health needs. Among individuals with frailty or complex multimorbidity, higher attainment of generic QIs was linked to decreased odds of outpatient non-attendance, while higher attainment of multimorbidity-specific QIs was associated with fewer outpatient visits.

### Comparison with existing literature

Our findings indicate that higher attainment of QIs is generally associated with increased planned secondary care use. Considered at its most straightforward level, higher QI achievement may be taken to indicate more active management of patients, and more active engagement of those patients with healthcare; for many individuals with multiple long-term conditions, this increased management /and engagement will include more outpatient appointments ^18^. Managing multimorbidity often necessitates multidisciplinary consultation, further contributing to the number of planned outpatient appointments. Notably, our results differed by generic and multimorbidity-specific QIs, especially for elective admissions.

Contrasting with previous evidence ^12,13^, we found that higher attainment of QIs was associated with increased use of unplanned care. The elevated likelihood of such healthcare use may not indicate care failure but rather may reflect successful risk detection through QI engagement or the inherent complexity of managing multimorbidity ^19^. Enhanced QI performance could promote early diagnoses, contributing to uncovering more unmet needs that subsequently drive greater secondary care use, including unplanned services. Furthermore, patients with higher QI attainment tend to be more clinically complex and frailer, having a greater risk of experiencing sudden acute events even with well-managed disease compared with healthy individuals.

Research on the effects of primary care interventions on hospital admissions shows mixed results ^20,21^. When considering interventions to reduce service utilisation more generally, those that measure patient-reported outcomes among patients with physical-mental comorbidity were more likely to demonstrate successful reductions in service utilisation ^22^. For example, chronic disease management in the community can reduce unplanned hospitalisations and ED visits ^23^. Some unplanned visits to emergency departments may be avoidable with tailored community-based interventions ^24^. A recent systematic review pointed out that community-based programs using a personalised approach and shared decision-making with the patient appeared to demonstrate a higher margin of effectiveness in reducing emergency department attendances ^25^. This highlights the need for holistic, patient-centred approaches beyond traditional QI metrics.

The severity of multimorbidity (e.g. frailty or number of comorbidities) appears to modify the association between primary care quality and secondary healthcare use, although the modifying effect varied across types of healthcare. For example, effect sizes of high multimorbidity-specific QIs for increasing ED visits were lower among non-frail patients than those with frailty. Frail patients with multimorbidity often have more comprehensive health needs, which may lead to greater needs in disease management such as urgent or unpredictable health events ^26^. High attainment of QIs is critical to address these complex health needs of frail individuals and prevent adverse health events (e.g. falls, fractures, etc.), though personalised support may still need to further attenuate the impacts of frailty. Our findings also indicate that within the complex multimorbidity cohort, patients with high multimorbidity QI attainment had lower odds of outpatient visits. Patients with complex multimorbidity face more challenges in managing multiple conditions that likely interact. High attainment of multimorbidity-specific QIs could support better disease control among patients, leading to fewer referrals and improved self-management for the high-need population.

### Strengths and limitations

The QI measures assessed in this study reflect active management processes, which captured regular care plan reviews, mobility and functional assessments, and screenings for mental health conditions, aligning with national guidelines ^27^. We adopted an evidence-based approach to measure QI using the SNOMED CT clinical codes, accounting for insights from our team of multimorbidity patients and clinicians. A key strength of this study was using a large EHR database, which provides a unique and reliable resource to measure and understand quality of primary care as documented by the primary care team. We made a composite of the individual items of QIs using principal components analysis to reduce the dimensions for analysis. However, like other analyses of composite measures, we cannot rule out some information loss, although all assumptions have been examined. This study involved a wide range of chronic health conditions relevant to multimorbidity, but it did not consider the cluster of multimorbidity patients. Further study could examine the performance of quality of primary care and its impact on health demands of secondary care in different patient groups beyond frailty, which could potentially identify those for whom care most needs to be improved. Further, the variations in the lagged effects of QI attainments on health demands of secondary care may be influenced by the time window of QI attainments and subsequent healthcare use, as well as the frequency of engagement with individual indicators. Patients with complex multimorbidity have more health demands for health checks in primary care, and they inherently have a higher risk of complications and thus more secondary care use. The associations between QIs and secondary care use may partly reflect underlying illness burden, although we adjusted for social and health-related confounders. Our QI measures focused on the management of multimorbidity and medicines, rather than whole-system care coordination, which would benefit from further study.

### Implications for research, policy and practice

This analysis provides insights into the relationship between attainment of quality indicators in primary care and secondary healthcare use, focusing on patients with multimorbidity. We did not find a clear pattern of secondary care use according to QI engagement; to clarify this, it may be necessary to involve quantitative indicators measuring patient perspectives ^28^, which can help capture the unmeasured experiences to support enhanced quality of primary care holistically. Further research could also explore facilitators and barriers to understand the reasons for different engagement levels of multimorbidity-specific QIs. This would advance understanding of the practical benefit of the guidelines and standards in the diagnosis, monitoring, and management of multimorbidity ^29^. Our findings highlight the potential associations between multimorbidity-specific QIs and reduced elective hospital admissions for patients with multimorbidity. A comprehensive management strategy, as reflected by multimorbidity-specific QIs, which involve more detailed and in-depth checks, could potentially better support disease management and prevent serious outcomes that lead to hospitalisation. However, given the positive association between multimorbidity QIs and unplanned hospitalisation, some healthcare emergencies for multimorbidity might be unavoidable or require more tailored clinical management planning than QIs alone. It may also be attributable to attainment of generic QIs, which were associated with better attendance at outpatient appointments. This could be explained by improved trust in care providers and empowering patients for shared disease management ^30^. Future research may need to follow up on the long-term impacts of quality of primary care on demands for secondary health and adverse health outcomes.

### Conclusions

Our study demonstrates that different aspects of primary care quality show varied associations with secondary care use among patients with multimorbidity. While we found no consistent association between higher overall quality and reduced hospital utilisation, attainment of multimorbidity-specific quality indicators was linked to fewer elective admissions. These findings underscore the complexity of managing multimorbidity and highlight the need to explore broader process measures of care quality. Future research should incorporate clinician and patient perspectives to better understand how engagement with quality indicators shapes long-term patterns of secondary care use.

## Declarations

### Ethics approval

The project proposal received approval from the WSIC SDRAG committee. The Discover research platform received ethical approval from the UK Health Research Authority (reference 23/WM/0196).

### Contributors

QG conceived and designed the study, acquired and verified the data, conducted data analyses, and wrote the draft of the manuscript. AB and BH contributed to study design, conceptualisation, data verification and analyses. All authors contributed to the interpretation and revision of the manuscript. All authors reviewed and approved the final manuscript.

### Data sharing

The Discover-NOW Whole System Integrated Care dataset (WSIC) was available through monitored request and approved by the Northwest London Data Access Committee (DAC) (https://discover-now.co.uk/).

## Data Availability

The Discover-NOW Whole System Integrated Care dataset (WSIC) was available through monitored request and approved by the Northwest London Data Access Committee (DAC).

## Acknowledgements

The study is funded by the NIHR Imperial Biomedical Research Centre and the NIHR Applied Research Collaboration Northwest London. ALN and PA are also funded by the NIHR North West London Patient Safety Research Collaboration. The funding bodies had no role in the study design, data analysis and interpretation, decision to publish, or manuscript preparation. The views expressed are those of the authors and not necessarily those of the affiliated institutions or the NIHR and the Department of Health and Social Care.

